# Proposing the Roommate Sleep Preference Questionnaire (ROOMPREF) with a free online roommate matching tool

**DOI:** 10.64898/2026.03.02.26347046

**Authors:** Matthew W. Driller, Haresh Suppiah

## Abstract

Shared sleeping quarters are commonplace in contexts such as athletes at major sporting events, academic dormitories, and military barracks, yet mismatched sleep preferences can undermine rest and ultimately, human behaviour and performance. We introduce the Roommate Sleep Preference Questionnaire (ROOMPREF), a brief eight-question survey capturing preferences for noise, lighting, and temperature tolerances, snoring behaviour, and chronotype. Responses feed into a free, web-based clustering tool built in Python, which flags preference conflicts, and implements adaptive K-Means clustering within sex–chronotype subgroups. A post-cluster swapping algorithm further mitigates residual mismatches, enhancing the room-matching process. The resource includes distribution charts, group summaries, and optional automated room allocations, with downloadable CSV outputs. We demonstrate its application in a pilot cohort, highlighting its potential to improve sleep outcomes across various use-cases. This free resource has the potential to alleviate mismatched rooming partners, resulting in enhanced sleep and wellbeing outcomes.

## Introduction

The dynamics of shared living spaces, whether in athletic teams, college dormitories, high school boarding schools, military barracks, or workplace accommodations, can significantly influence sleep quality and, consequently, wellbeing and performance. Sleep, an indispensable component of human health, is intricately tied to our internal biological clocks and personal sleep preferences. When individuals with divergent sleep patterns cohabit, the potential for sleep disturbances increases (1, 2), thereby underscoring the importance of understanding and accommodating these differences for optimal coexistence. The challenge of maintaining optimal sleep conditions is magnified in shared accommodation, where individual preferences and habits can vary widely. During major events like the Olympic and Paralympic Games for example, athletes, who often dedicate their entire careers to training for these events, may have to endure sharing small rooms in the Athlete Village. During the 2016 Olympic Games in Rio, interventions aimed at improving sleep quality for elite swimmers led to significant enhancements in total sleep time and reductions in sleep latency, ultimately benefiting their performance, particularly for those reaching the finals (3). This study illustrates the direct link between sleep management strategies and athletic success, emphasising the need for tailored sleep support systems in shared lodging situations.

Similar sleep-related stressors are also well documented in other group-living settings. In military contexts, irregular sleep schedules, mismatched routines, and shared spaces have been linked to poorer sleep quality and reduced alertness (4, 5). Likewise, in educational settings such as college dormitories or boarding schools, conflicting chronotypes, differing bedtime behaviors, and preferences among roommates can result in increased sleep disturbances and decreased academic performance (6-8). These findings underscore the need for structured roommate-matching strategies that consider individual sleep traits.

To address this, we propose a brief questionnaire designed for use across diverse shared accommodation contexts. In combination with a freely accessible online clustering tool, coordinators, whether team managers, residential staff, or dormitory advisors - can use the results to group individuals with compatible sleep profiles. Matching is based on two main domains: chronotypes and sleep environment preferences. With this information, coordinators can then match roommates based on their own knowledge of other factors (e.g., personality types), or they can opt for the tool to automatically allocate rooms. This tool supports more restful sleep and improved coexistence in any setting where individuals must share sleeping quarters.

## Materials and Methods

### Proposing the ROOMPREF

The Roommate Sleep Preference Questionnaire (ROOMPREF) that we propose (see supplementary files), may be a crucial tool in navigating the complexities of shared living situations. By addressing the multifaceted nature of sleep and circadian rhythms, this questionnaire facilitates a deeper understanding and appreciation among roommates of their respective sleep needs. As evidenced by the cited studies, aligning sleep environments and habits with individual preferences and biological rhythms is paramount in fostering a healthy, productive, and harmonious shared living experience. This approach underscores the necessity of accommodating individual sleep characteristics to enhance communal living dynamics and overall well-being, and human performance. The proposed questionnaire below incorporates key features that we believe are important to room-sharing; chronotypes and sleep environment preferences and are based on previously reported factors that may differ among individuals, resulting in conflicts (9-12).

### Free online tool

We have developed a free online tool for administrators, coordinators, practitioners, and/or managers to use when allocating rooms. First, we provide a Google Form template for managers/coordinators to collect the ROOMPREF Questionnaire, then an app-based tool for analysis and allocation of roommates. The tool is housed here: https://www.siestaresearch.com/roompref

### Tool development

The ROOMPREF app is a custom-built roommate compatibility algorithm designed to assist with optimal room assignments in shared sleep environments. The app stratifies participants by sex, accounts for chronotype compatibility, and clusters individuals based on sleep environment preferences. It includes conflict-prevention logic (e.g., avoiding pairing snorers with participants who require silence, where possible), adaptive clustering logic to prevent impractical room sizes, and filters to exclude incompatible chronotype pairings (e.g., morning and evening types in the same room).

### Participants and Data Collection

First, participants complete the ROOMPREF questionnaire in the provided Google Form. The complete ROOMPREF questionnaire is included below, however, it takes less than 2 minutes to complete, and includes the following:

- Sex (or preferred roommate sex)
- Noise sensitivity during sleep
- Lighting and temperature preferences
- Snoring behavior
- Chronotype (morning-evening preference)
- Optional open-text comments (e.g., can indicate preferred roommate)

All data are downloaded from Google Forms as a CSV format and uploaded into the ROOMPREF web application.

### Data Processing and Workflow

1. Gender-Based Stratification Participants are first grouped by their response to Sex (or preferred roommate sex), ensuring that clustering occurred only within gender-appropriate groups.
2. Chronotype Classification Chronotype answers are mapped into three categories via a simple lookup:

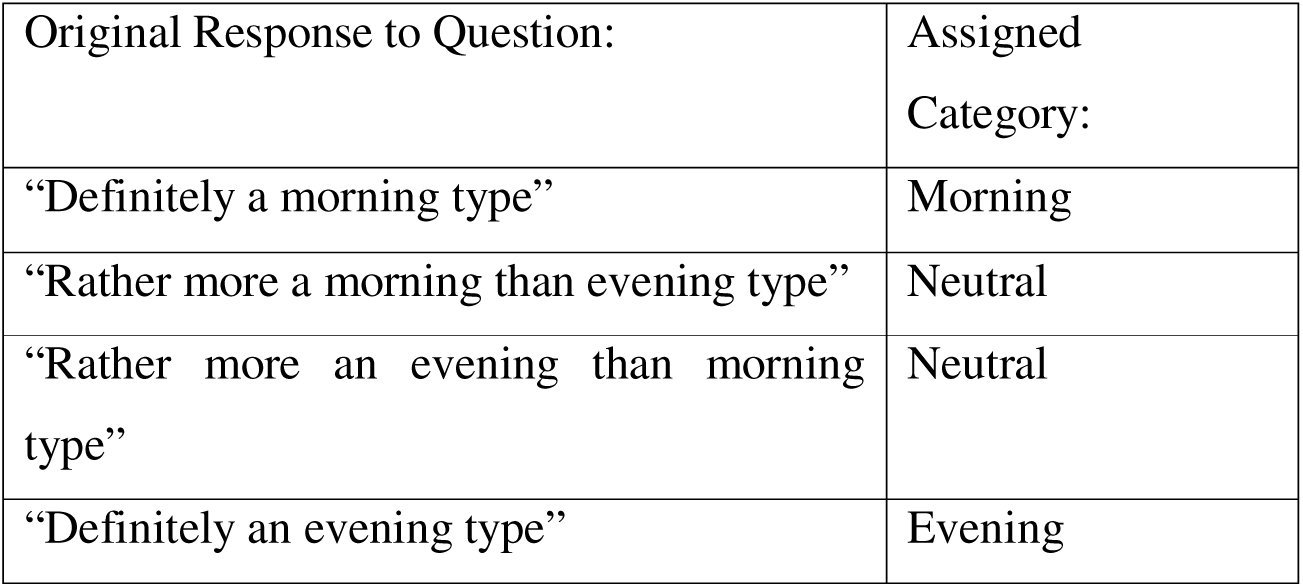 Participants are grouped accordingly for compatibility evaluation.
3. Conflict Exclusion Logic Two exclusions are enforced before clustering:

- Snoring conflicts: If a participant reported “Yes” to snoring and another in the subgroup required “Absolute Silence”, both are excluded from clustering together (unless absolutely necessary, in which case a “warning” will be flagged).
- Chronotype mismatch: After initial clustering, any resulting rooms containing both “Morning” and “Evening” types are filtered out to prevent incompatible circadian preferences (unless absolutely necessary, in which case a “warning” will be flagged).
4. Clustering Variables and Feature Encoding Clustering is based on:

- Noise sensitivity
- Lighting preference
- Temperature preference Each feature is label-encoded, and z-score standardised. Chronotype is not used in the clustering itself but instead constrained during final room allocation.
5. Adaptive Clustering Strategy K-Means clustering is applied within each gender–chronotype subgroup:

- Groups with ≤3 participants: assigned to one cluster (k=1)
- Groups with ≥4 participants: split into two clusters (k=2)

The adaptive clustering strategy (k=1 for N≤3; k=2 for N≥4) employs a heuristic approach to k-selection. This method prioritises the creation of practically sized initial clusters suitable for typical room allocation scenarios (e.g., forming pairs from a group of 4 or 5) and prevents excessive fragmentation of smaller subgroups. While this approach does not optimise k based on intrinsic data structure (e.g., via silhouette analysis), it aligns with the tool’s primary goal of providing coordinators with a manageable and interpretable starting point for room assignments.

### Room Swapping Algorithm (when automatic room allocation is triggered)

- If a conflict remains, we search among the other rooms in the same cluster for a “tolerant” roommate (i.e. someone whose noise tolerance is not Absolute Silence).
- We then swap the silence sensitive participant in the conflicted room with the tolerant participant from the other room.
- This operation is done in a round-robin fashion so that no room ends up exceeding the chosen room size.

By appending this swap logic after standard clustering and before final export, we ensure that, wherever possible, snorers are relocated into rooms with roommates who can tolerate ambient noise, while still respecting room size constraints and never leaving anyone unassigned.

### Output and Visualisation

The app produces:

- A table of final room assignments (with warnings where relevant)
- Flags for conflicts such as snorer + silence-sensitive pairings, incompatible chronotypes, and underfilled rooms
- A downloadable CSV matching the visual output
- A toggle option to “enable automatic roommate allocation” based on selected room size (2, 3, or 4).

### Implementation

The app was built using:

- Python for logic and data processing
- pandas for data manipulation
- scikit-learn for clustering
- streamlit for the web interface

The app supports CSV uploads, previewing, automatic or manual allocation toggling, conflict filtering, and export of final group assignments.

## Results

### Use case example

In the lead up to the 2024 Paralympic Games in Paris, we ran a pilot trial of the ROOMPREF questionnaire and roommate compatibility clustering app with 51 athletes (21 female, 30 male). In the Athlete Village, it was two people per room. Previously, roommates were matched largely on athletes’ preference, which could still be taken into account by indicating this in the comments section of the survey. The example below (Figure 1) shows the workflow that was implemented.

**Figure 1.**
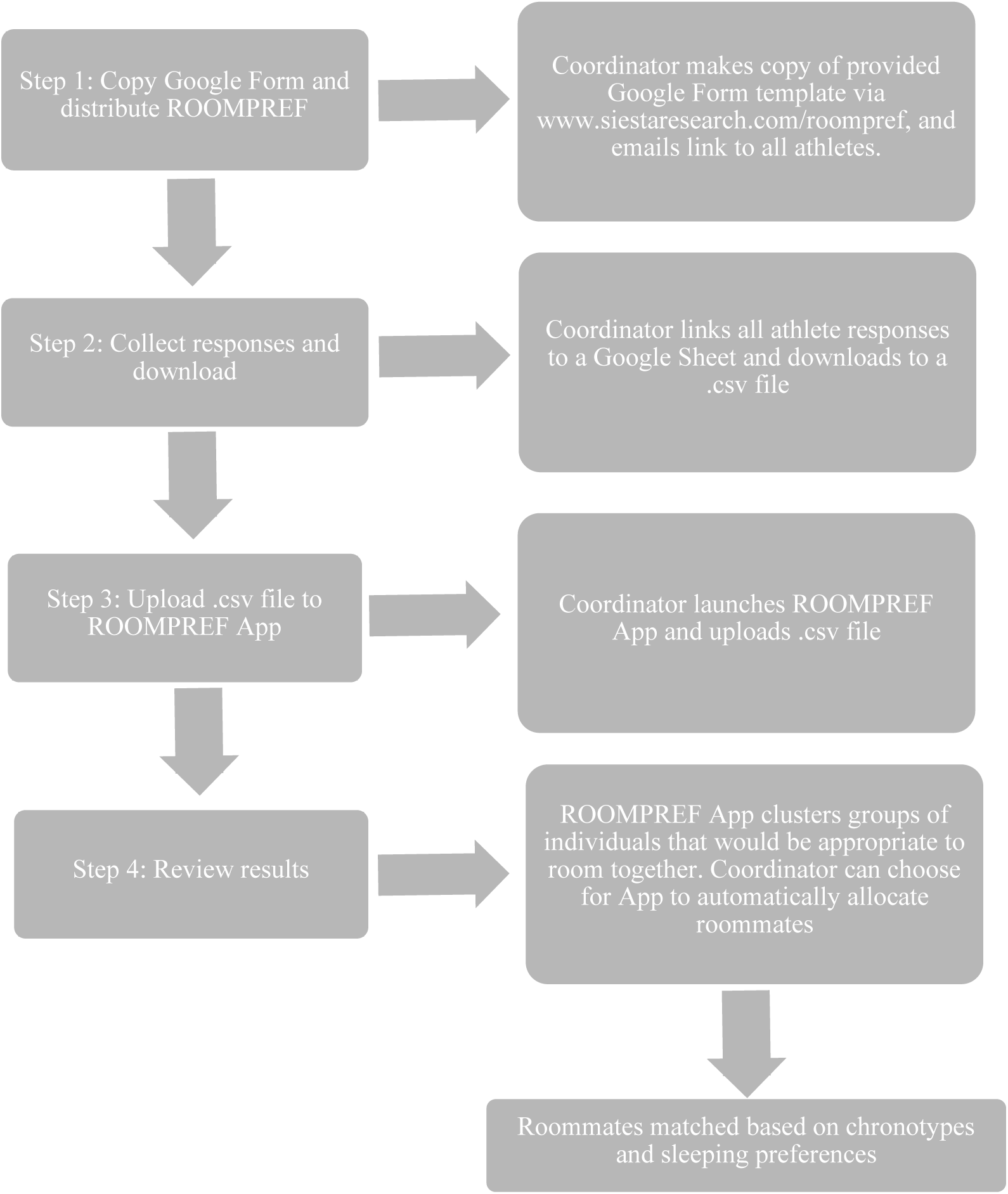
Workflow of using the ROOMPREF roommate matching tool.

Once the coordinator had collated all results, and uploaded the file to the ROOMPREF App, they were be presented with the results, first with a summary of chronotype distribution, noise, lighting, and temperature distribution from respondents (Figure 2), then with a participant grouping summary, placing individuals in groups that would be appropriate to room with each other (Figure 3). If the coordinator chose to have rooms automatically allocated (see further details in methods), they were presented with a further table of roommate matching based on room size (Figure 4). Data screenshots below are from the real-life scenario presented above, but with individual names changed to fictional characters to preserve anonymity.

**Figure 2.**
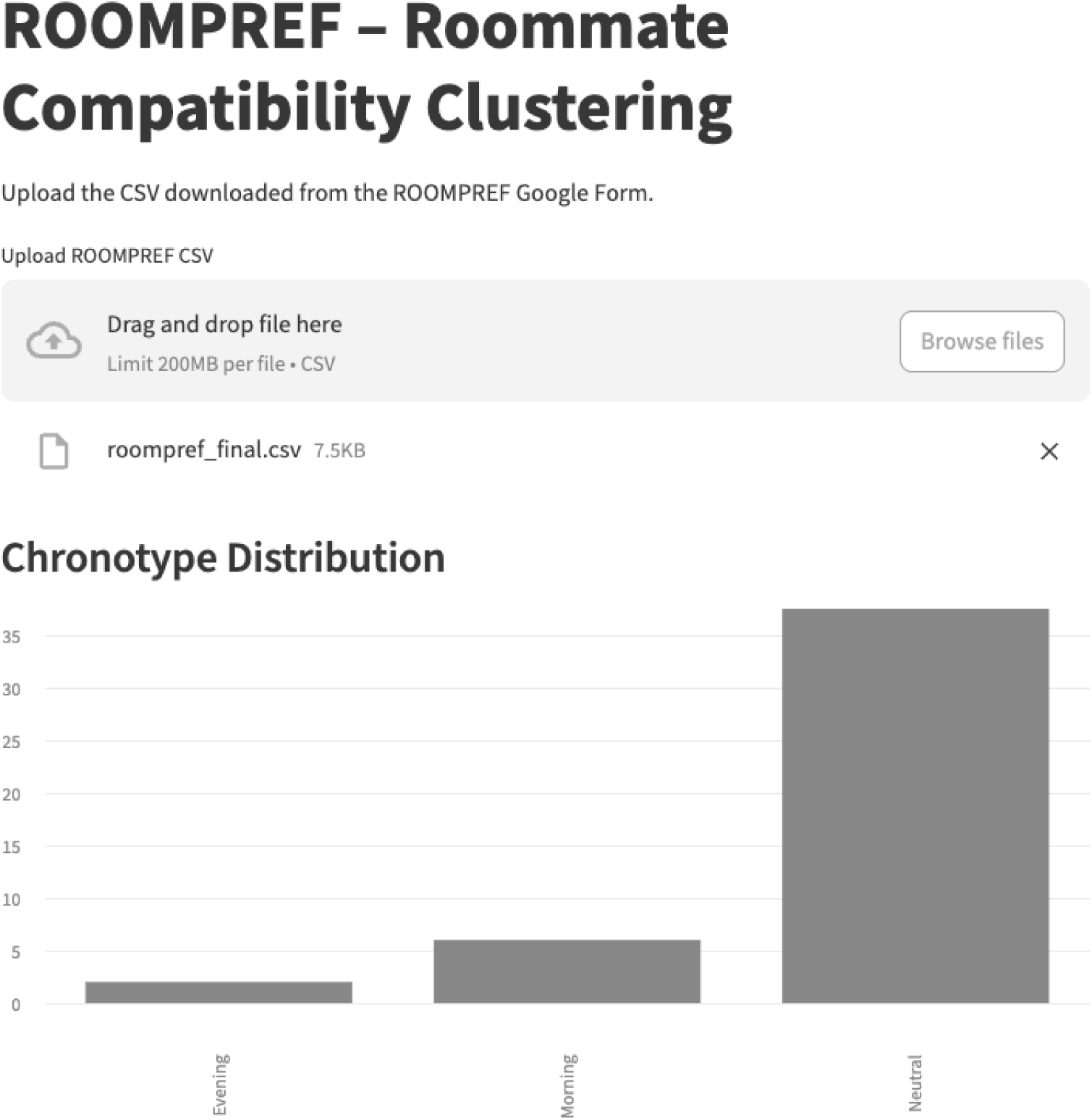
On uploading the CSV file of collected results, coordinators are presented with some summary figures of distributions across chronotypes, noise, lighting, and temperature preferences.

**Figure 3.**
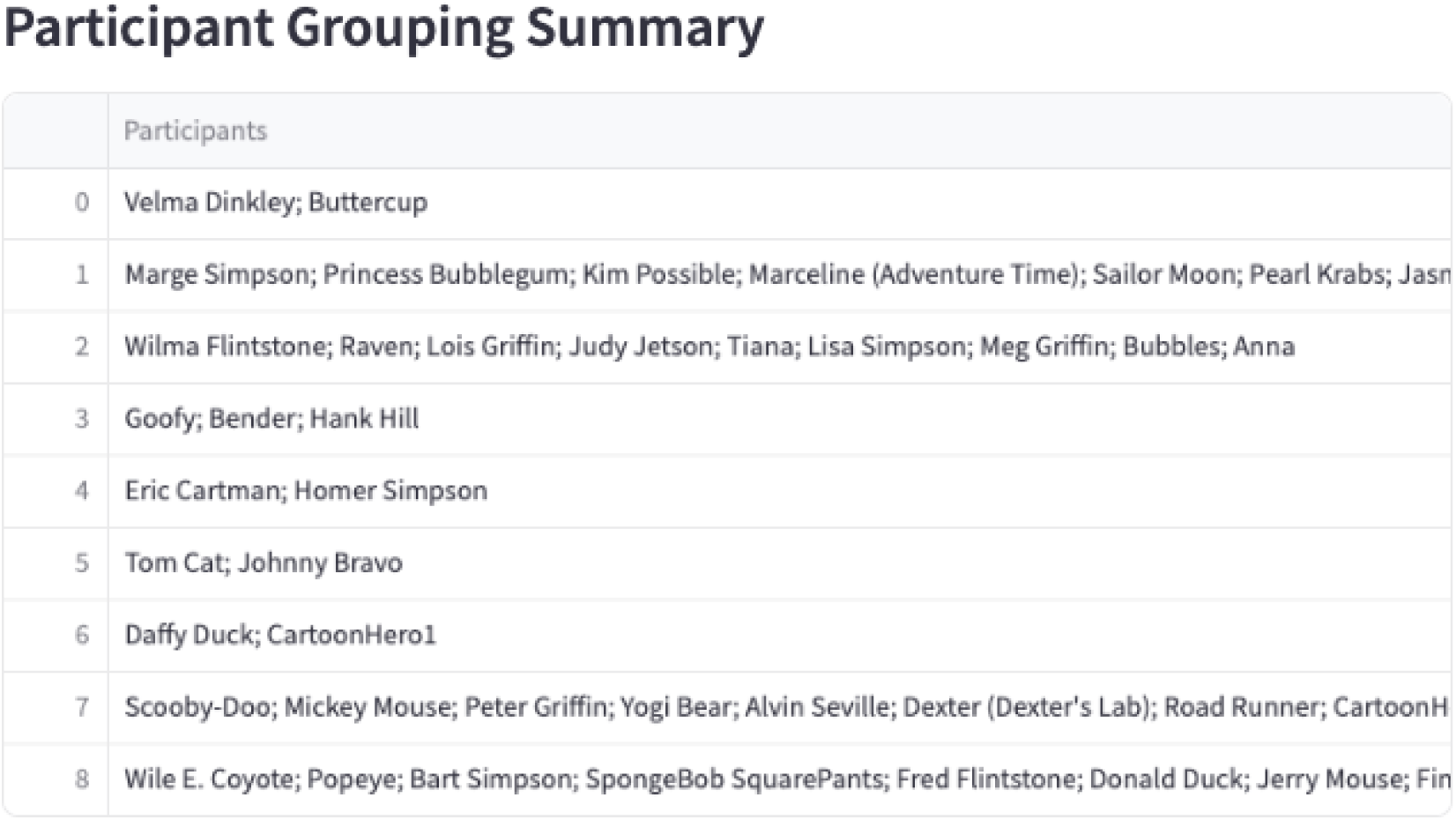
Coordinators will be provided with a list of groups with individuals that would be appropriate to room together based on chronotypes and sleep preferences. They can then choose to allocate themselves from within those groups (e.g., using knowledge of personalities etc.), or choose to automatically allocate based on room size.

**Figure 4.**
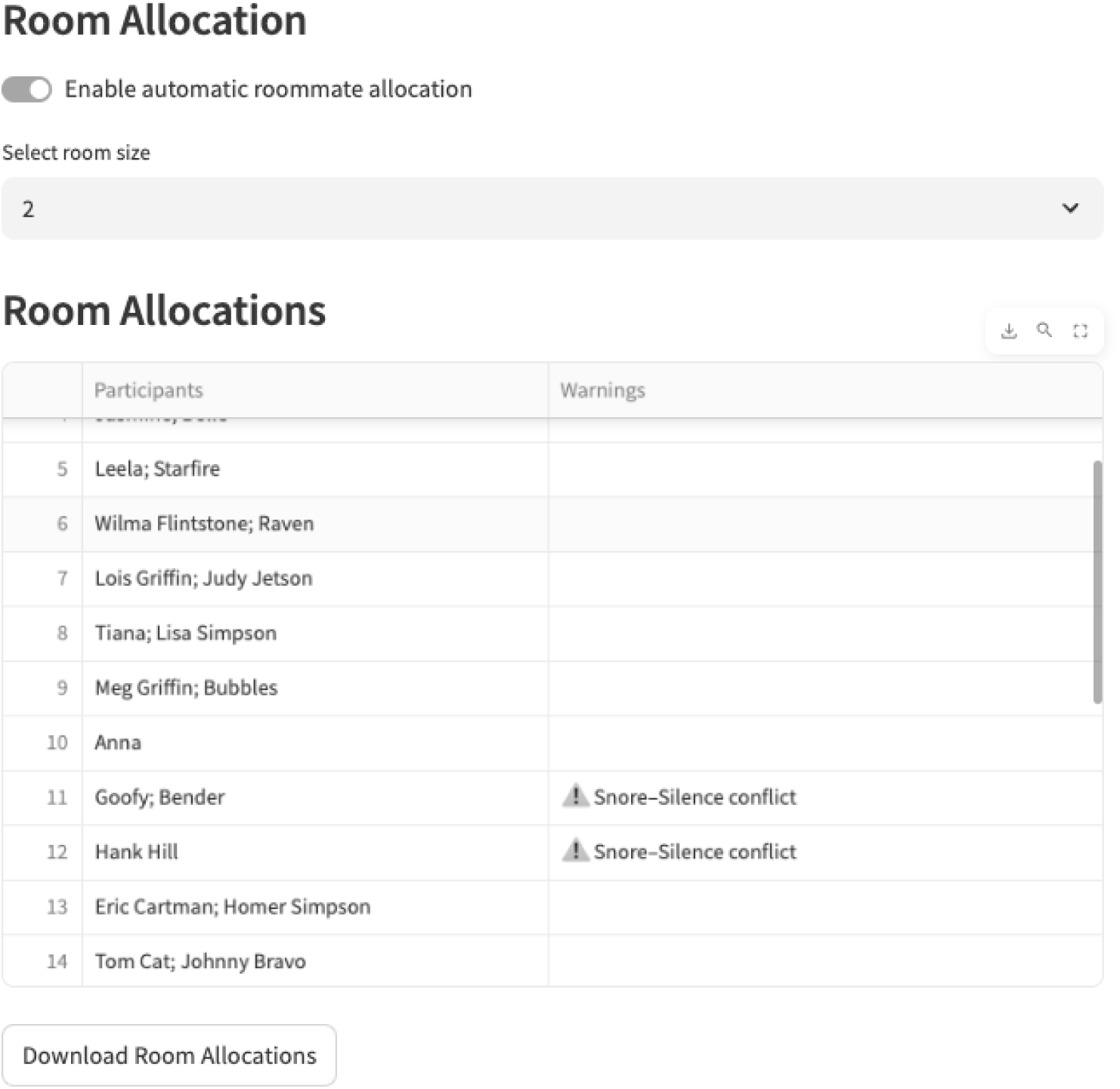
In this example, if the coordinator chooses to automatically allocate rooms based on 2 per room, they will be provided with the list of roommate matches. Warnings will be provided for any conflicts that exist (e.g., Snore-Silence conflict), and some rooms may be underfilled if they are leftover from the matching process (as seen above). Coordinators then have the option to download room allocations as a CSV file.

## Discussion

In this paper we introduce the Roommate Sleep Preference Questionnaire (ROOMPREF) and a free, web-based clustering tool designed to support compatibility-based room allocations in settings where shared sleeping quarters are unavoidable. Rather than testing a specific intervention on sleep outcomes, the primary contribution is methodological: a brief, operationally practical questionnaire focused on key sleep-related preferences, coupled with a transparent, rule-based clustering algorithm that can be implemented at scale by coordinators with minimal technical expertise. The worked example from a sporting team at a major competition demonstrates the feasibility of integrating this workflow into real-world operations, from survey distribution through to automatic room allocation and exportable room lists, without the need for specialised programming skills.

The need for such a tool is underscored by existing evidence that room configuration and co sleeping arrangements meaningfully influence sleep. A study by Costa et al. (13) investigated the impact of different sleeping arrangements on sleep in thirteen elite male youth soccer players during an official training camp. Athletes who slept in individual bedrooms obtained longer and more efficient sleep, as well as shorter sleep latency, compared to those sharing rooms (13). While individual rooms are clearly the best case scenario, this is often unrealistic due to financial or logistical constraints. For example, providing single rooms for all athletes at a Paralympic Games would substantially increase village size and cost; for team sports that travel regularly, room sharing is often the only cost effective option. ROOMPREF is therefore intended as a pragmatic strategy to reduce obvious mismatches within the constraints of shared accommodation, not as an alternative to single rooms where those are feasible.

Beyond elite sport, multiple group living environments face analogous challenges. In residential education settings such as college dormitories and boarding schools, students are frequently assigned roommates with minimal attention to sleep preferences or habits, despite evidence that poor or irregular sleep is associated with impaired academic performance and wellbeing (6-8, 14-16). Similarly, military personnel often report high rates of sleep disorders, with shared sleeping environments, irregular schedules and environmental disturbances identified as important contributors to inadequate sleep and reduced operational readiness (4, 5, 12). ROOMPREF is intentionally generic and freely accessible so that coordinators in schools, universities, military units and other group living contexts can use the same questionnaire and clustering logic to bring greater structure and defensibility to room allocation decisions.

A distinctive feature of ROOMPREF is the explicit integration of chronotype information into the allocation workflow. Circadian misalignment and mismatched chronotypes are linked to poorer health, cognitive performance and academic outcomes (8, 14, 17-19). Within the ROOMPREF app, chronotype responses are mapped into three categories (morning, neutral and evening), and clustering occurs within sex–chronotype subgroups, with subsequent logic to avoid placing Morning and Evening types in the same room wherever possible. This design choice reflects the growing recognition that circadian factors are not merely theoretical constructs but have tangible implications for day to day functioning, especially in athletes, students and military personnel who must perform at specific times of day. Although the current paper does not evaluate downstream sleep or performance outcomes, the tool operationalises this evidence by embedding chronotype compatibility into the default rooming algorithm.

The questionnaire and tool also emphasise environmental sleep preferences (noise, lighting and temperature), which are consistently identified as key determinants of sleep quality and next day functioning (9-12, 20). ROOMPREF encodes these preferences and applies several layers of conflict prevention logic; for example, snorers are flagged and, where possible, are not paired with individuals who indicate a need for absolute silence, and remaining conflicts are addressed via a post cluster room swapping algorithm that searches for “tolerant” roommates within the same cluster. This approach does not guarantee perfect matching but provides a reproducible, auditable framework that is substantially more structured than informal manual allocation and can be refined as more implementation data accumulate.

From a methodological standpoint, the tool prioritises usability and interpretability over algorithmic complexity. The adaptive K means strategy (k = 1 for very small groups, k = 2 for larger subgroups) is a pragmatic heuristic tailored to typical room sizes (two to four occupants) and to the realities of coordinators who need quickly interpretable groupings rather than opaque optimisation outputs. More sophisticated approaches (for example, data driven selection of k, probabilistic or multi objective optimisation) could, in theory, refine allocations further, but may reduce transparency and usability for end users. The present implementation should therefore be viewed as a practical starting point that can be iteratively improved as organisations gain experience and additional data on roommate satisfaction and sleep outcomes.

The pilot application with Paralympic athletes highlights several operational strengths of the ROOMPREF workflow. It shows that the end to end pipeline - from distributing the questionnaire via an online form, exporting responses as a CSV file, uploading to the web app, and generating room assignments with conflict flags and downloadable outputs, can be executed within existing organisational practices and technology platforms. Importantly, the tool is designed to complement, not replace, coordinator judgement: users can inspect clustered groups that would be appropriate to room together and either manually assign rooms based on additional knowledge (for example, personality, team dynamics or accessibility needs) or enable automatic allocation with clear warnings for any residual conflicts.

Several limitations should be acknowledged. First, this paper primarily describes the development and workflow of the questionnaire and tool rather than providing a full validation of their impact on sleep, wellbeing or performance. We did not collect objective or subjective sleep data pre and post implementation, nor did we quantify roommate satisfaction or perceptions of fairness in room allocations. As such, the benefits of ROOMPREF remain theoretically grounded rather than empirically confirmed. Second, the questionnaire itself has not yet undergone psychometric evaluation (for example, reliability, factor structure and sensitivity to change). Although it was intentionally designed to be brief and face valid, future work should assess whether the items capture distinct, stable dimensions of sleep related preferences and whether additional questions (for example, bedtime routines, technology use or health conditions) would meaningfully improve matching. Third, the current implementation relies on self reported preferences and behaviours, which may be imperfect or influenced by social desirability (for example, under reporting snoring). Misreporting could lead to suboptimal matches despite the best efforts of the algorithm. Fourth, the allocation logic focuses on a limited set of variables and does not currently incorporate broader psychosocial factors such as personality compatibility, interpersonal history or cultural considerations, all of which may influence roommate harmony. ROOMPREF is therefore best viewed as a tool for systematically aligning key sleep related factors, to be used alongside, not instead of, local knowledge and consultation with the individuals involved.

Future research should move beyond descriptive demonstration to formal evaluation of the tool’s impact. Randomised or quasi experimental studies could compare standard room allocation procedures with ROOMPREF guided allocation in athletes, boarding students or military units, examining sleep outcomes (for example, actigraphy and sleep diaries), daytime functioning, performance and roommate satisfaction. At the methods level, expanding the algorithm to incorporate additional preference dimensions, exploring alternative clustering approaches, and enabling iterative reallocation (for example, for longer term dormitory placements) are logical extensions. There is also scope to integrate ROOMPREF into broader digital ecosystems (such as team management platforms or student housing systems) and to co design versions tailored to specific populations, including para athletes or individuals with particular medical or accessibility needs.

In summary, ROOMPREF and its accompanying free online clustering tool offer a practical, scalable approach to bringing sleep science into the everyday logistics of shared accommodation. By explicitly capturing chronotype and core sleep environment preferences, and embedding them in an easily deployable algorithm, this resource provides coordinators with a structured way to reduce obvious mismatches in shared rooms. The next step is to rigorously test whether such compatibility based allocation translates into better sleep, wellbeing and performance across the diverse settings in which people are required to share their sleeping space.

## Data Availability

All data produced in the present study are available upon reasonable request to the authors

## Availability of data and materials

Online tool and code provided via https://www.siestaresearch.com/roompref

## Code availability

https://github.com/drillz77/roompref-clustering

