## Appendices for "Proposing the Roommate Sleep Preference Questionnaire (ROOMPREF) with a free online roommate matching tool"

**Appendices – ROOMPREF**

**ROOMPREF: Roommate Sleep Preference Questionnaire**

**Basic Information:**

1. Name:

2. Sex (if you are non-binary, or prefer not to disclose, what sex would you prefer to room with?):

- [ ] Male

- [ ] Female

3. What level of noise can you tolerate while sleeping?

- [ ] Absolute Silence

- [ ] Low Background Noise

- [ ] Doesn't Matter

4. Preferred lighting conditions for sleeping:

- [ ] Complete Darkness

- [ ] Dim Light

- [ ] Doesn’t Matter

5. Preferred room temperature for sleeping:

- [ ] Cool (≤ 20°C, 68°F)

- [ ] Moderate (21-24°C, 70 - 75°F)

- [ ] Warm (≥ 25°C, 77°F)

6. Do you snore or have been told that you snore?

- [ ] Yes

- [ ] No

7. One hears about “morning” and “evening” types of people. Which one of these types do you consider yourself to be?

- Definitely a “morning” type

- Rather more a “morning” than “evening” type

- Rather more an “evening” than “morning” type

- Definitely an “evening” type

**Additional Comments:**

8. Any other information or preferences you would like to share about your sleeping habits or needs? Indicate here if you have a preferred roommate.
